# Molecular and serological investigation of the 2021 COVID-19 case surge in Mongolian vaccinees

**DOI:** 10.1101/2021.08.11.21261915

**Authors:** Naranjargal J. Dashdorj, Naranbaatar D. Dashdorj, Mitali Mishra, Lisa Danzig, Thomas Briese, W. Ian Lipkin, Nischay Mishra

**Affiliations:** Onom Foundation, Ulaanbaatar, Mongolia; Center for Infection and Immunity, Columbia University, New York, USA; PANDEFENSE Advisory, USA

## Abstract

A surge in Covid-19 cases in Mongolia in March 2021 resulted in a government-mandated shutdown. This shutdown was relaxed in May 2021 as case numbers decreased and nationwide vaccination rates using Sinopharm, Covishield/AstraZeneca, Sputnik V, and Pfizer/BioNTech vaccines exceeded 50% of the population. Case rates increased again in early June 2021 in both vaccinated and non-vaccinated individuals. To determine whether the surge was due to the emergence of Delta or another variant, or vaccine failure, a rapid, opportunistic investigation was conducted that comprised virus sequence analysis of nasal swab samples from breakthrough cases and antibody assays of plasma from healthy vaccinees. More than 90% of breakthrough infections during the second case surge were due to the Alpha variant. Spike protein ELISA and SARS-CoV-2 neutralization assays data revealed large differences in plasma titers of antibodies to SARS-CoV-2, with mRNA vaccines eliciting higher titers than adenovirus-vectored or killed virus vaccines.

## Introduction

In March 2021 an increase in COVID-19 cases in Mongolia (population 3.3 million) led to a government-mandated shutdown and a nationwide vaccination program with four different vaccines. In early June, a reduction in cases attributed to vaccination led to relaxation of the shutdown that was followed by a surge in disease (Figure. 1).

**Figure 1.**
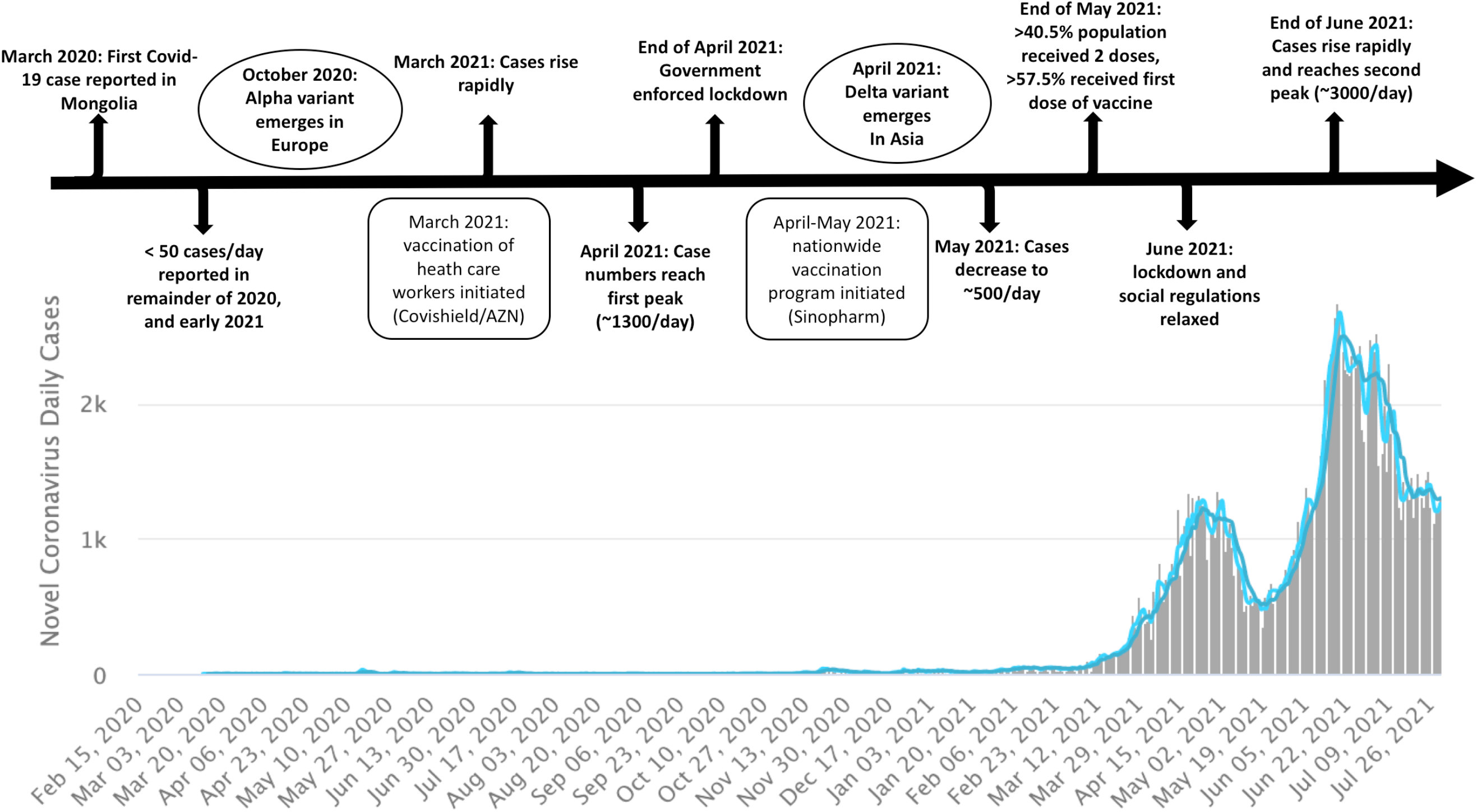
**A]** Time line of COVID-19 in Mongolia February 2020 - July 2021; **B]** Illustration of daily new cases in Mongolia with 3 days and seven days moving average of daily new cases (blue plot lines) Source: https://www.worldometers.info/coronavirus/country/mongolia/

To determine whether the surge was due to the emergence of Delta or another variant, or vaccine failure, a rapid, opportunistic investigation was conducted that comprised virus sequence analysis of nasal swab samples from breakthrough cases and antibody assays of plasma from healthy vaccinees.

## Methods

Samples were collected with informed consent. RNA from nasal swabs was extracted for Illumina sequencing using the Mybaits capture system (Arbor Biosciences). Reads were mapped and aligned to SARS-CoV-2 reference sequence (accession no. NC_045512) and variant analyses performed using GISAID, NextClade[1]. Plasma from healthy vaccinees were screened for anti-SARS-CoV-2 antibodies in EUROIMMUN Anti-SARS-CoV-2 IgG ELISA and EUROIMMUN Anti-SARS-CoV-2 NCP IgG ELISA. Neutralizing antibodies (NAb) to Washington, and Delta SARS-CoV-2 variants were quantitated in an absolute end point neutralization assay that measures 100% neutralizing activity with a SARS-CoV-2 qPCR [2].

## Results

We sequenced 97 nasal samples from Covid-19 patients with breakthrough infections after vaccination (Supplementary Table 1), and tested plasma from 100 healthy subjects for antibodies to SARS-CoV-2 (surface glycoprotein, S and nucleocapsid phosphoprotein, NCP), and NAb after Covishield/AstraZeneca, Sputnik V, Sinopharm, or Pfizer/BioNTech vaccination (Table 1).

**Table 1:** SARS-CoV-2 immunoreactivity of plasma samples from 100 healthy vaccinated individuals; as determined by S-ELISA, NCP-ELISA, and end point absolute viral neutralization assay against WA strain and Delta variant. Samples were collected with informed consent under the auspices of Onom IRB 241 and IRB 242.

| Serological assay | Status | Pfizer/BioNTech (n=24) | Covishield/AZN (n=24) | Sputnik V (n=24) | Sinopharm (n=24) | Sinopharm + Pfiz/BioN* (n=4) | chi-square test significant (<0.05) p values (total positive vs negative) |
| --- | --- | --- | --- | --- | --- | --- | --- |
| Spike ELISA | Negative | 0 (0.00%) | 5 (20.83%) | 12 (50%) | 17 (70.83%) | 0 (0.00%) | Pfizer/BioNTech vs Sputnik V=0.0002, Pfizer/BioNTech vs Sinopharm=0.0001, Covishield/AZN vs Sputnik V=0.03, Covishield/AZN vs Sinopharm=0.0005 |
|  | Total positive | 24 (100.00) | 19 (79.17%) | 12 (50%) | 7 (29.17%) | 4 (100%) |  |
|  | Weak positive | 2 (8.33%) | 2 (8.33%) | 5 (20.83%) | 4 (16.67%) | 1 (25.00%) |  |
|  | Positive | 22 (91.67%) | 17 (70.83%) | 7 (29.17%) | 3 (12.5%) | 3 (75.00%) |  |
| NCP ELISA | Negative | 23 (95.83%) | 21 (87.50%) | 23 (95.83%) | 21 (87.5%) | 3 (75.00%) | NA |
|  | Total positive | 1 (4.17%) | 3 (12.50%) | 1 (4.17%) | 3 (12.5%) | 1 (25.00%) |  |
|  | Weak positive | 1 (4.17%) | 2 (8.33%) | 0 (0.00%) | 0 (0.00%) | 0 (0.00%) |  |
|  | Positive | 0 (0.00%) | 1 (4.17%) | 1 (4.17%) | 3 (12.5%) | 1 (25.00%) |  |
| Neutralizing Ab (WA) | Not detected | 5 (20.83%) | 12 (50.0%) | 18 (75.00%) | 19 (79.17%) | 1 (25.00%) | Pfizer/BioNTech vs Covishield/AZN=0.03, Pfizer/BioNTech vs Sputnik V=0.0002, Pfizer/BioNTech vs Sinopharm=0.00005, Covishield/AZN vs Sinopharm=0.03 |
|  | Detected | 19 (79.17%) | 12 (50.0%) | 6 (25.00%) | 5 (20.83%) | 3 (75.00%) |  |
|  | ≤1:40 | 14 (58.30%) | 4 (16.70%) | 4 (16.70%) | 2 (8.33%) | 2 (50.00%) |  |
|  | ≥1:40 | 5 (20.83%) | 8 (33.30%) | 2 (8.33%) | 3 (12.50%) | 1 (25.00%) |  |
| Neutralizing Ab (Delta) | Not detected | 17 (70.83%) | 15 (62.50%) | 22 (91.6%) | 21 (87.5%) | 3 (75.00%) | Covishield/AZN vs Sputnik V=0.016, Covishield/AZN vs Sinopharm=0.045 |
|  | Detected | 7 (29.17%) | 9 (37.50%) | 2 (8.33%) | 3 (12.5%) | 1 (25.00%) |  |
|  | ≤1:40 | 4 (16.67%) | 3 (12.50%) | 1 (4.20%) | 2 (8.33%) | 0 (0.00%) |  |
|  | ≥1:40 | 3 (12.50%) | 6 (25.00%) | 1 (4.20%) | 1 (4.20%) | 1 (25.00%) |  |

\*Sinopharm + Pfiz/BioN group was not included in statistical analysis due to small sample size. NA- No significant (<0.05) p value was recorded for NCP ELISA.

Full-length SARS-CoV-2 sequences were obtained from 92 samples. Eighty-seven of 92 (94.6%) were Alpha variant. Five of 92 (5.4%) were Delta variant. Of 87 Alpha variant breakthrough infections, 54 (62.1 %) were from Sinopharm, 7 (8.0%) from Sputnik V, 23 (26.4%) from Covishield/AZN, and 3 (3.4%) from Pfizer/BioNTech vaccinees. Of 5 breakthrough Delta variant infections, 1 (20%) was from Sinopharm, 3 (60%) were from Covishield/AZN, and 1 (20%) was from Pfizer/BioNTech vaccinees (Supplementary Table 1).

More subjects had spike ELISA reactivity after Pfizer/BioNTech (91.6%), and Covishield/AstraZeneca (70.8%), than Sputnik V (29.2%) or Sinopharm (12.5%). Nucleoprotein reactivity was present in 12.5% of Sinopharm vaccinees. One Covishield/AstraZeneca vaccinee was NP positive, presumably because of infection. Only Sinopharm contains NP. Three of four subjects (75%) who received two Sinopharm doses followed by one Pfizer/BioNTech dose were spike-positive (Table 1) (Supplemental Figure 1). More subjects had NAb to WA strain after Pfizer/BioNTech (79.7%), and Covishield/AstraZeneca (50.0%), than after Sputnik V (25.0%) or Sinopharm (20.8%). Neutralizing antibodies to Delta variants were present in fewer subjects after Pfizer/BioNTech (29.2%), Sputnik V (8.33%), and Sinopharm (12.5%). NAb to Delta variants were detected in nine subjects after Covishield/AZN (37.50%); however, 3/9 were NCP ELISA-positive indicating previous SARS-CoV-2 exposure. Eliminating these three subjects would decrease the percentage with NAb to Delta to 25%. Three of four vaccinees who received a single dose of Pfizer/BioNTech after two doses of Sinopharm had NAb to WA; one had NAb to Delta.

## Discussion

Our findings provide additional support for ordinal hierarchy of protection amongst vaccine platforms, with mRNA vaccines performing better than adenovirus-vectored or killed virus vaccines [3]. They also confirm that current vaccines, based on the prototypic WA strain, elicit only minimal levels of NAb to current circulating strains [4 5]. The case surge in Mongolia in June 2021 did not reflect the introduction of the Delta variant but premature relaxation of physical distancing and masking. These findings underscore the importance of continuous surveillance for virus evolution, coordinated modification of vaccines, and a cautious approach to the elimination of classical social controls for mitigating contagion.

## Limitations

Our study was limited in numbers and in its retrospective collection of specimens.

## Data Availability

Data is available on request.

## Article Information

### Corresponding Authors

Nischay Mishra, PhD, ND Dashdorj, and W. Ian Lipkin, MD

### Author Contributions

Dr. Mishra & Dr. Lipkin had full access to all of the data in the study and takes responsibility for the integrity of the data and the accuracy of the data analysis.

#### Concept and design

Mishra, Lipkin, Danzig, Nara Dashdorj, and ND Dashdorj

#### Acquisition, analysis, or interpretation of data

All authors.

#### Drafting of the manuscript

Mishra, Lipkin

#### Critical revision of the manuscript for important intellectual content

All authors.

#### Statistical analysis

Mishra, Lipkin

#### Obtained funding

Lipkin

#### Administrative, technical, or material support

Mishra, Lipkin, Nara Dashdorj, and ND Dashdorj

#### Supervision

Mishra, Briese, Lipkin

### Additional Contributions

Nancy Messonnier; MD (Skoll Foundation) and Larry Brilliant; MD(Pandefense) provided insights into study design. We are grateful to CII staff members; Komal Jain; MS for bioinformatics analyses, Lokendra Chauhan;MS and Eliza Choi;MS for neutralization assays, Riddhi Thakkar;BS for ELISA assays, James Ng;BS for RNA extractions and qPCRs, Alexandra Oleynik Petrosov; MS and Alper Gokden; MS for high throughput sequencing, and Pryanka Sharma;MS for sample shipment and logistics. We would like to thank Sumiya Byambabaatar; MS, Purevjargal Bat-Ulzii, Anir Enkhbat, Enkhtuul Batbold, Delgersaikhan Zulkhuu, Byambasuren Ochirsum, Tungalag Khurelsukh, Ganbold Dalantai, Natsagdorj Burged, Uurtsaikh Baatarsuren, Nomin Ariungerel, Odgerel Oidovsambuu, Andreas S. Bungert, Zulkhuu Genden, Dahgwahdorj Yagaanbuyant, and Altankhuu Mordorj for sample collection and administrative support.

