## Supplemental appendix for "Molecular and serological investigation of the 2021 COVID-19 case surge in Mongolian vaccinees"

### Samples

*Nasal swab samples from subjects with breakthrough infections (n=97)*

Samples were collected with informed consent under the auspices of Onom IRB 241 and IRB 242. We obtained nasal swabs from 97 subjects with breakthrough infections after vaccination. Fifty-two (53.6%) were females (average age: 42.8±13.8 years); 45 (48.4%) were males (average age: 41.9±15.5 years). Eighty-four (86.6%) were fully vaccinated i.e. received both (prime and booster) doses of vaccines; 13 (13.4%) received only the first (prime) dose of vaccine prior to breakthrough infection. Out of 84 who received both doses, 53 (63.1%) received Sinopharm, 28 (33.3%) received Covishield/AstraZeneca, 2 (2.4%) received Sputnik, and one (1.2%) received Pfizer/BioNTech.

| **Vaccine name** | **Sex (M/F) (subject numbers) (n=97)** | **Age in years (average ± S.D.)** | **Two doses vaccinee (n=) vs single dose vaccinee (n=)** | **Interval between second vaccine dose and infection (average days ± S.D.)** | **Interval between only vaccine dose and infection (average days ± S.D.)** | **Total SARS-CoV-2 complete genome sequence recovered (n=92)** | **No. of Alpha variant (n=87)** | **No. of Delta variant (n=5)** |
|---|---|---|---|---|---|---|---|---|
| Sinopharm | M (27) | 47.07±15.86 | 25 vs 2 | 40.32±20.6 | 43.50±11.3 | | | |
| | F (30) | 45.8 ±13.53 | 28 vs 2 | 45.58±21.5 | 43.00±16.0 | | | |
| | Total (57) | 46.4 ±14.69 | 53 vs 2 | 43.06±20.6 | 43.25±11.3 | 55 | 54 | 1 |
| Covishield | M (13) | 32.3 ±10.92 | 13 vs 0 | 66.85±5.2 | NA | | | |
| | F (15) | 39.67±12.64 | 15 vs 0 | 68.38±5.2 | NA | | | |
| | Total (28) | 36.25±12.43 | 28 vs 0 | 67.79±53.5 | NA | 26 | 23 | 3 |
| Sputnik V | M (4) | 36.75±10.54 | 0 vs 4 | NA | 50.75±20.2 | | | |
| | F (4) | 41.75±10.73 | 2 vs 2 | 87.00±7.00 | 36.5±0.5 | | | |
| | Total (8) | 39.25±10.92 | 2 vs 6 | 87.00±7.00 | 46±17.8 | 7 | 7 | 0 |
| Pfizer BioNTech | M (1) | 49.00 | 1 vs 0 | 7.00±0 | NA | | | |
| | F (3) | 29.3±1.24 | 0 vs 3 | NA | 25.73±19.7 | | | |
| | Total (4) | 34.25±8.58 | 1 vs 3 | 7.00±0 | 25.33±19.7 | 4 | 3 | 1 |

Supplemental table 1- Demographic information of Covid-19 patients with breakthrough infections

*Plasma samples from healthy vaccinated individuals (n=100)*

Samples were collected with informed consent under the auspices of Onom IRB 241 and IRB 242. Of 100 subjects, 24 (24%) received two doses of Sinopharm, 24 (24%) received two doses of Covishield/AZN, 24 (24%) received two doses of Sputnik, 24 (24%) two doses of Pfizer/BioNTech (prime and booster); 4 (4%) received two doses of Sinopharm and one dose of Pfizer/BioNTech. All samples were tested for anti-spike SARS-CoV-2 antibodies, and anti-nucleocapsid-phosphoprotein SARS-CoV-2 antibodies using

EUROIMMUN ELISA kits, and for neutralizing antibodies using an end-point viral neutralization assay [1].

### SARS-CoV-2 Sequencing

RNA extracted using QIAcube HT (Qiagen) was employed for SARS-CoV-2 complete genomic sequencing. Illumina libraries were prepared and enriched for SARS-CoV-2 sequences using Mybaits capture system (Arbor Biosciences) [2]. Captured libraries were pooled and sequenced on the Illumina Nextseq 2000; and ~6-8 million reads/samples were generated. After demultiplexing, reads were mapped and aligned against the SARS-CoV-2 reference sequence (accession no. NC_045512).

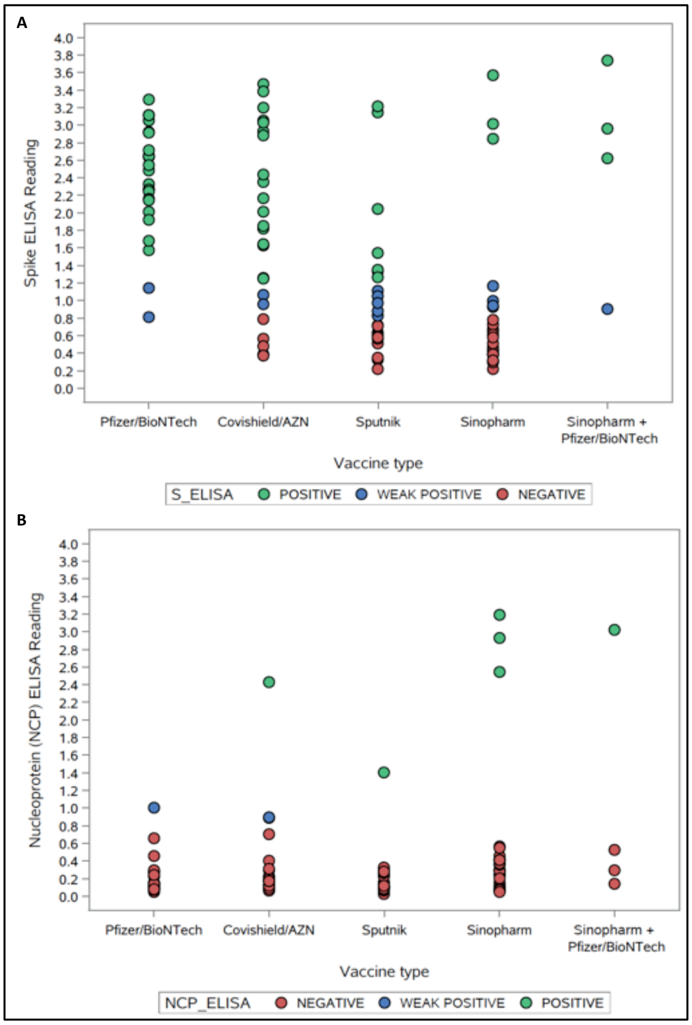


**Supplemental Figure 1.** A] Scatter plot showing EUROIMMUN Anti-SARS-CoV-2 IgG ELISA readings optical density (O.D.); B] Scatter plot showing EUROIMMUN Anti-SARS-CoV-2 NCP IgG ELISA readings (O.D.). Negative < 0.8 O.D.; Weak positive 0.8-1.1 O.D.; Positive >1.1 O.D.

1. Eckhardt CM, Cummings MJ, Rajagopalan KN, et al. Evaluating the efficacy and safety of human anti-SARS-CoV-2 convalescent plasma in severely ill adults with COVID-19: A structured summary of a study protocol for a randomized controlled trial. Trials 2020;**21**(1):499 doi: 10.1186/s13063-020-04422-y[published Online First: Epub Date]|.
2. Tillett RL, Sevinsky JR, Hartley PD, et al. Genomic evidence for reinfection with SARS-CoV-2: a case study. Lancet Infect Dis 2021;**21**(1):52-58 doi: 10.1016/S1473-3099(20)30764-7[published Online First: Epub Date]|.
